# Patterns of Glucose Metabolism in [^18^F]FDG PET Indicate Regional Variability and Neurodegeneration in the Progression of Alzheimer’s Dementia

**DOI:** 10.1101/2023.11.10.23298396

**Authors:** John J. Lee, Tom Earnest, Sung Min Ha, Abdalla Bani, Deydeep Kothapalli, Peiwang Liu, Aristeidis Sotiras, the Alzheimer’s Disease Neuroimaging Initiative

**Affiliations:** Mallinckrodt Institute of Radiology, Washington University, Saint Louis, Missouri, USA; Neuroimaging Laboratory and Research Center, Washington University, Saint Louis, Missouri, USA; Computational Imaging Research Center, Washington University, Saint Louis, Missouri, USA; Institute for Informatics, Data Sciences and Biostatistics, Washington University, Saint Louis, Missouri, USA

**Keywords:** neurodegeneration, glucose metabolism, fluorodeoxyglucose, positron emission tomography, nonnegative matrix factorization, Alzheimer’s, dementia

## Abstract

In disorders of cognitive impairment, such as Alzheimer’s disease, neurodegeneration is the final common pathway of disease progression. Modulating, reversing, or preventing disease progression is a clinical imperative most likely to succeed following accurate and explanatory understanding of neurodegeneration, requiring enhanced consistency with quantitative measurements and expanded interpretability of complex data. The on-going study of neurodegeneration has robustly demonstrated the advantages of accumulating large amounts of clinical data that include neuroimaging, motiving multi-center studies such as the Alzheimer’s Disease Neuroimaging Initiative (ADNI). Demonstrative advantages also arise from highly multivariate analysis methods, and this work reports advances provided by non-negative matrix factorization (NMF). NMF revealed patterns of covariance for glucose metabolism, estimated by positron emission tomography of [^18^F]fluorodeoxyglucose, in 243 healthy normal participants of ADNI. Patterns for glucose metabolism provided cross-sectional inferences for 860 total participants of ADNI with and without cerebral amyloidosis and clinical dementia ratings (CDR) ranging 0-3. Patterns for glucose metabolism were distinct in number and topography from patterns identified in previous studies of structural MRI. They were also distinct from well-establish topographies of resting-state neuronal networks mapped by functional magnetic resonance imaging. Patterns for glucose metabolism identified significant topographical landmarks relating age, sex, APOE ε4 alleles, amyloidosis, CDR, and neurodegeneration. Patterns involving insular and orbitofrontal cortices, as well as midline regions of frontal and parietal lobes demonstrated the greatest neurodegeneration with progressive Alzheimer’s dementia. A single pattern for the lateral parietal and posterior superior temporal cortices demonstrated preserved glucose metabolism for all diagnostic groups, including Alzheimer’s dementia. Patterns correlated significantly with topical terms from the Neurosynth platform, thereby providing semantic representations for patterns such as attention, memory, language, fear/reward, movement and motor planning. In summary, NMF is a data-driven, principled, supervised statistical learning method that provides interpretable patterns of neurodegeneration. These patterns can help inform the understanding and treatment of Alzheimer’s disease.

**Highlights:**

▪ Data-driven non-negative matrix factorization (NMF) identified 24 canonical patterns of spatial covariance of cerebral glucose metabolism. The training data comprised healthy older participants (CDR = 0 without amyloidosis) cross-sectionally drawn from ADNI.
▪ In *healthy* participants, mean SUVRs for specific patterns in precuneus, lateral parietal cortex, and subcortical areas including superficial white matter and striatum, demonstrated increasing glucose metabolism with advancing age.
▪ In *asymptomatic participants with amyloidosis*, glucose metabolism increased compared to those who were *asymptomatic without amyloid*, particularly in medial prefrontal cortex, frontoparietal cortex, occipital white, and posterior cerebellar regions.
▪ In *symptomatic participants with amyloidosis*, insular cortex, medial frontal cortex, and prefrontal cortex demonstrated the most severe losses of glucose metabolism with increasing CDR. Lateral parietal and posterior superior temporal cortices retained glucose metabolism even for CDR > 0.5.
▪ NMF models of glucose metabolism are consistent with models arising from principal components, or eigenbrains, while adding additional regional interpretability.
▪ NMF patterns correlated with regions catalogued in Neurosynth. Following corrections for spatial autocorrelations, NMF patterns revealed meta-analytic identifications of patterns with Neurosynth topics of fear/reward, attention, memory, language, and movement with motor planning. Patterns varied with degrees of cognitive impairment.

## Introduction

Over the lifespan, the human brain undergoes organizational development into young adulthood, then continues organizational processes with increasing variability into late adulthood. Alzheimer’s disease may be regarded as a disease of microscale proteinopathies and macroscale alterations of brain networks^1^. By progressive neurodegeneration processes, functional connectivity networks degrade, and cognitive functions deteriorate. The types of networks that fail determine characteristic dementia phenotypes, generating variability of presentations, even with the presence of indistinguishable proteinopathies. Nevertheless, phenotypes can associate with specific lesions to brain networks and subnetworks^2^. Neurodegeneration is a state-trajectory with varieties of cognitive and disease outcomes. Typical Alzheimer’s disease has pathophysiology leading to a progressive amnestic syndrome involving the default mode network^3^, but atypical Alzheimer’s pathophysiology can involve non-memory systems such as language, vision, and executive systems^4,5^.

This work follows prior computational models demonstrating that data from Alzheimer’s disease can yield predictive features in amyloid data, tau data, and the final common pathway of neurodegeneration^6^. Integrative computational models for clinical symptoms, degenerative brain anatomy in [^18^F]fluorodeoxyglucose positron emission tomography (FDG PET), and refined considerations of functional brain networks have previously by reported^2^. Integrative computational models for Alzheimer’s disease should incorporate “large-scale ensembles of coordinated neuronal activity” and “large-scale network topologies”^1,2^. These are motivated by prior results for the default mode network and failing of interactions with other brain network hubs^3,7,8^. Disease and neurodegeneration are disruptions of functional networks, but other complex characteristics necessarily contribute: molecular processes, microscale misfolding of proteins, mesoscale functional operations.

Organizational features of the brain can be informed by previously established methodologies. Cognitive ontologies such as perception, emotion, memory, social cognition, language, executive function, and their neuroanatomical localization have been described, and ontological correspondences can be constructed with tools such as Neurosynth^9,10^. Evolutionary expansion in early development has previously been observed through structural magnetic resonance imaging (MRI), cortical thickness estimations with FreeSurfer^11^, and identification of patterns of covariance (PoC) using non-negative matrix factorization (NMF)^12^. NMF manages data complexity using multivariate bases, or patterns of covariance. NMF is distinguishable from principle components analysis (PCA) and independent component analysis (ICA); NMF patterns are sparse and compact, with interpretable parts^13^. This work demonstrates that NMF for FDG PET produces patterns of covariance distinct from those seen in structural NMF and functional connectivity. These features unique to FDG PET are likely to reflect the underlying modality as NMF itself remains hypothesis free, depending only on data, the numerical regularizations provided by nonnegativity, and linear factorization. FDG PET is an accepted biomarker for neurodegeneration^14^. Thereby, NMF can indicate regions specific for regional parts-based features that exhibit loss of glucose metabolism on FDG PET, thereby indicating neurodegeneration.

In all, lower-dimensional models suitable for characterizing Alzheimer’s disease are sought for their interpretability, and NMF can provide such models.

## Results

This study of participants in ADNI had cross-sectional design, selecting the earliest available clinical data and neuroimaging following participant enrollments. All data from ADNI had registry-confirmed exam and imaging dates between 2005 Dec 15 and 2020 Mar 4. PoC for FDG derived from 243 participants without cerebral amyloidosis and CDR=0, aged 56 – 95 (mean 73.5 ± std 6.5), 48.3% with female sex, 0 – 2 APOE ε4 alleles (mean 0.19 ± std 0.43). Analyses of neurodegeneration as estimated by FDG used four additional diagnostic cohorts, defined by CDR and presence or absence of amyloidosis, and detailed further in Table 1. Strategies for analyses depended on availability of FDG with contemporaneous T1-weighted MRI, which provided for spatial normalizations. Analyses also required availability of FreeSurfer-derived regions of interest (ROI) from pons and cerebellar vermis, as well as meta-ROIs^15^, for consistency of standardized uptake value ratios (SUVR) with existing reports drawn from ADNI. Separation of diagnostic cohorts required PET with amyloid-targeting tracers and CDR. Figure 1 illustrates additional details of data inclusion and exclusion.

**Figure 1.**
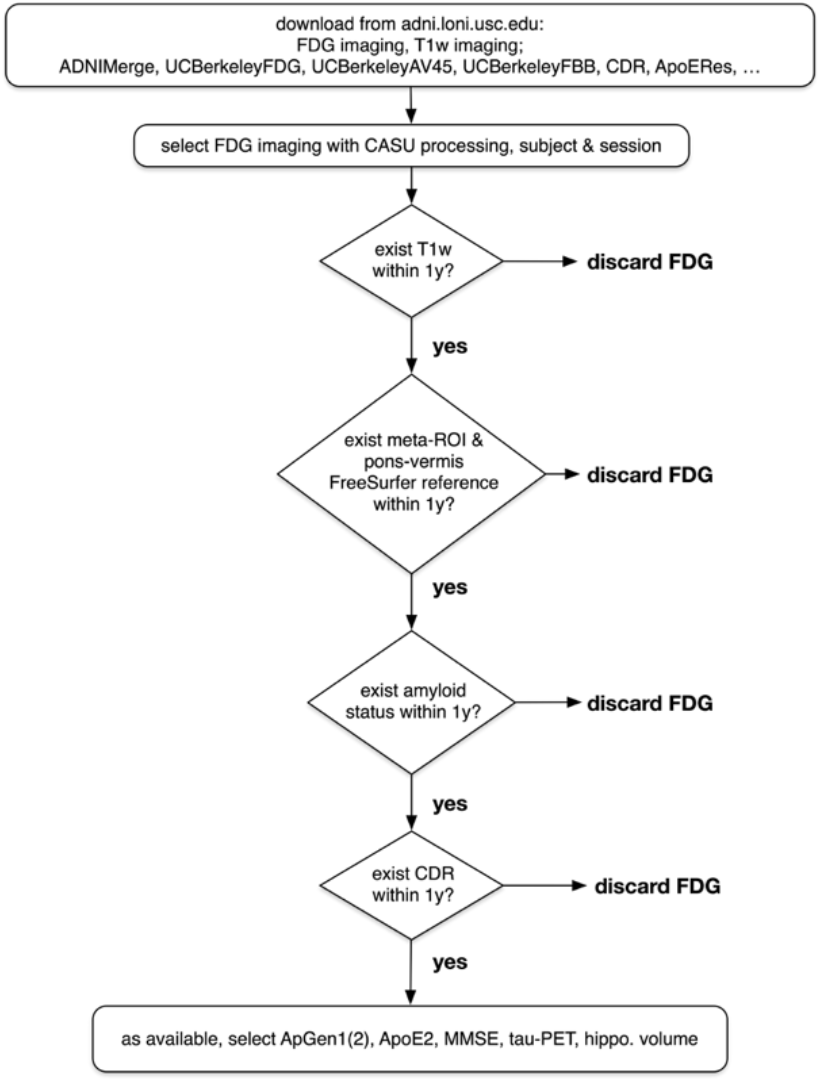
Inclusion an exclusion of data for analyses.

**Table 1.** Census of participants from ADNI. T1w scans and pons-vermis reference regions within 1 year separation are more informative for inferences. Author JJL made detailed visual inspections of registration quality of CDR=0 and amy-cases, excluding two FDG session for poor registration with the MNI150 atlas. The first available FDG session for each study participant provided cross-sectional inferences. Analyses of covariances used data with complete CDR, amyloid status, age, sex, APOE ε4 data, and valid pattern-weighted averages of imaging. Descriptive statistics (mean ± std. dev.) for age, sex, and ApoE4 inform 1^st^ FDG scans.

| Groups | CDR = 0, amy- | CDR = 0, amy+ | CDR=0.5, amy+ | CDR > 0.5, amy+ |
| --- | --- | --- | --- | --- |
| no. FDG with cross-sectionally complete covariates | 243 | 106 | 402 | 109 |
| Age range (years at enrollment) | 56 – 95 | 60 – 91 | 55 – 92 | 56 – 96 |
| Age (years at enrollment) | $73.5 \pm 6.5$ | $75.8 \pm 6.3$ | $74.3 \pm 7.4$ | $75.4 \pm 8.1$ |
| Female (%) | 48.3 | 62.3 | 44.8 | 52.4 |
| No. ApoE4 range (alleles) | 0 – 2 | 0 – 2 | 0 – 2 | 0 – 2 |
| No. ApoE4 (alleles) | $0.19 \pm 0.43$ | $0.53 \pm 0.57$ | $0.84 \pm 0.70$ | $0.95 \pm 0.66$ |

### NMF Identifies Metabolic Networks

Analysis of reconstruction error and split-half reproducibility provided model selection. The gradient of reconstruction error reached plateaus at 11, 18 and 24 patterns. Additional improvement in the gradient beyond 24 raised concerns for overfitting. Split-half reproducibility with 49 independent anticlustering splits favored 2, 12, and 24 patterns. The distributions of anticlustered adjusted Rand index are shown in Figure 2. Local maxima were discernable by distribution medians denoted by dashes.

**Figure 2.**
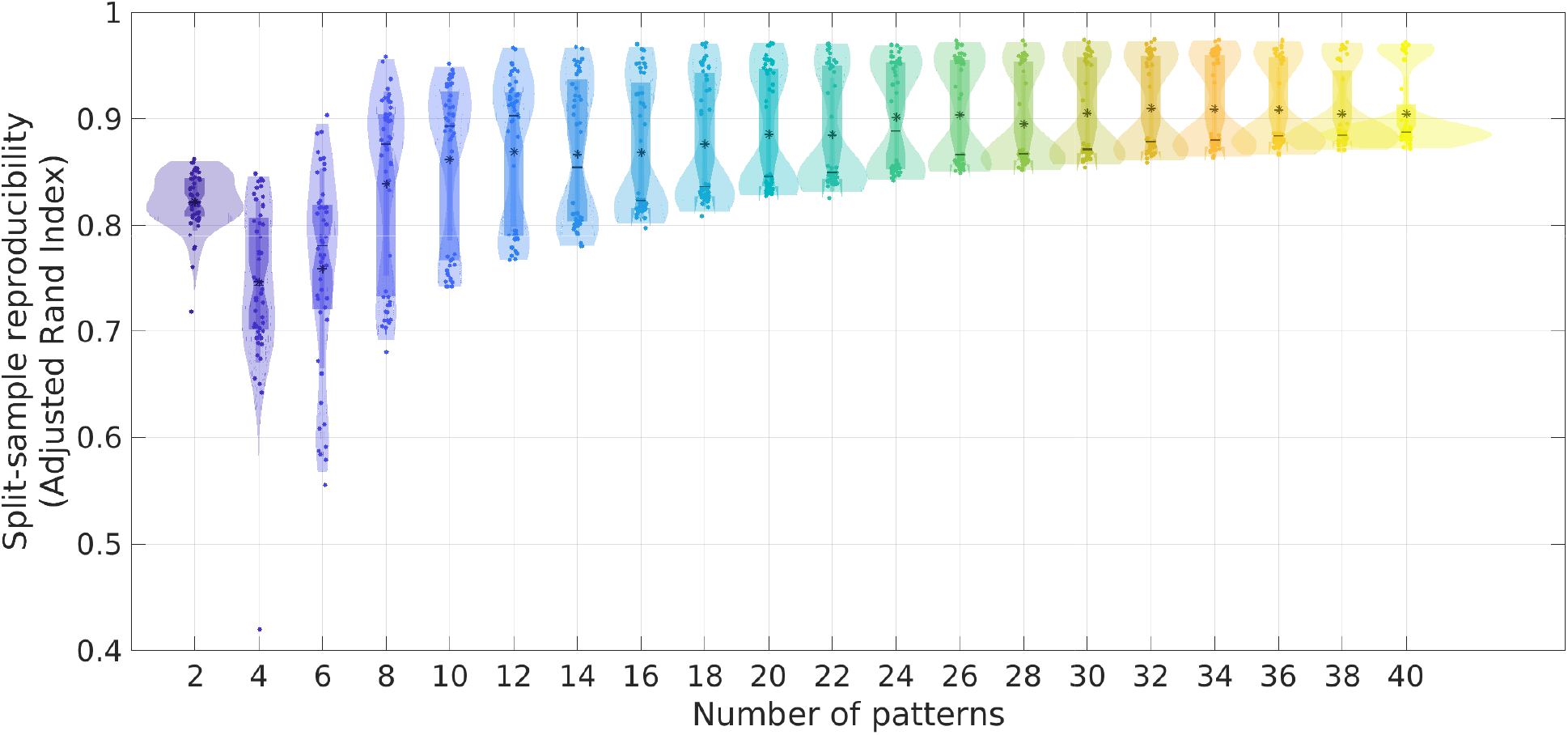
Split-sample reproducibility with anticlustering algorithms. 49 repetitions of anticlustering produced distributions of adjusted Rand index for each proposed model of NMF patterns of covariance: 2 - 40. Local peaks of adjusted Rand index included 2, 12, and 24.

Examination of models with 2, 12, and 24 patterns suggested hierarchical organization for some patterns. Two patterns supported separation into cortical regions that indicated intrinsic and extrinsic large-scale functional networks, originally noted in FDG PET^16^. Intrinsic networks match default mode and frontoparietal functionality observed in resting-state functional magnetic resonance imaging (fMRI)^17^. Extrinsic networks match the complementary resting-state networks, including somatomotor sensory, visual, auditory and attention networks^18^. This dichotomy has been replicated by alternative methods including estimates of myelination made from comparisons of T1 and T2 weighting^19^ and construction of generalized coordinates delineating the separation of primary motor and sensory cortices from heteromodal association cortices^20^. The dichotomy has also been replicated in NMF studies of cortical thickness and evolutionary expansion^12^, which associated patterns of the highest evolutionary areal expansion with topographies reproducibly identified with the default mode and frontoparietal control functional networks. Patterns of lower evolutionary areal expansion associated with topographies identified with visual cortex, somato-motor-sensory areas, auditory cortex, limbic structures, the insula with areas associated with the ventral attention functional network, and the dorsal attention network. The model of 12 patterns revealed further parcellations of intrinsic and extrinsic functional networks, but not according to familiar topographies. Notably, the default mode network segregated into distinct regions for the orbitofrontal and medial temporal poles, the medial prefrontal and limbic areas with striatum, and the lateral parietal areas with large confluences of the lateral frontal lobes. The centrum semiovale formed an independent pattern. The cerebellum and midbrain also formed an independent patter. Twenty-four patterns reproduced all patterns of the 12-pattern model, while introducing additional segregations, many of which corresponded to known cortical surface topographies and subcortical segmentations.

For the 24-pattern model, enumeration of pattern anatomy & correspondence with Brodmann areas (BA) are below. While NMF does not impose any ordering to its patterns of decomposition, distinct from the ordering of explained variance produced by principle component analysis (PCA), we imposed ordering of 24 enumerated patterns according to the pattern-averaged quantity of FDG scaled as SUVR. Thereby, pattern 1 had highest FDG SUVR, indicating maximal glucose metabolism, while pattern 24 had the least FDG SUVR.

Pattern 1: maximal FDG SUVR ranged across the lateral left hemisphere, encompassing lateral frontal areas, lateral parietal areas, and superior temporal areas. SUVR also encompassed middle frontal and inferior frontal gyri (BA45, BA44), inferior somato-motor-sensory regions (BA43), and extended along the supramarginal and angular gyri (BA40, BA39). SUVR also extended into posterior aspects of the superior temporal gyrus (BA22, BA42, BA41). Moderate SUVR localized to the lateral aspect of the right premotor cortex at its intersection with the middle frontal gyrus (BA6). Moderate SUVR also localized to the precuneus (medial BA7), without involving the superior parietal lobule. Minimal SUVR localized to the right putamen.

Pattern 2: maximal SUVR was bilateral, symmetric, and encompassed insular cortices (BA13), opercular parts of the inferior frontal gyri (BA45, BA44), and postcentral regions (BA43).

Moderate SUVR involved the medial prefrontal cortices (BA10, BA11). Moderate SUVR also localized to the caudate heads and anteromedial thalamus.

Pattern 3: maximal SUVR was bilateral, symmetric, and predominantly ranged across the medial surfaces of the frontal and parietal lobes, including the dorsolateral prefrontal cortices (BA9), frontal eye fields (BA8), premotor and supplementary motor cortices (BA6), primary somato-motor-sensory cortices (BA1-BA4), somatosensory association cortices (BA5), and precuneus (medial BA7). Tapering SUVR reached the superior margins of the dorsal cingulate cortices (BA24, BA31). Minimal SUVR localized to anterior and dorsal thalamus.

Pattern 4: maximal SUVR was bilateral, symmetric, and covered the temporal poles (BA38), extending into inferior temporal gyri (BA20, BA37). Moderate SUVR included entorhinal cortex (BA34, BA28), amygdala, and hippocampal structures (BA35).

Minimal SUVR localized to orbital inferior frontal gyri (BA47/12).

Pattern 5: maximal SUVR was bilateral, symmetric, and specifically localized to the grey-white junctional regions of the cerebellum. Moderate SUVR extended into the midbrain.

Pattern 6: maximal SUVR was bilateral, symmetric, and ranged along dorsolateral surfaces posterior to the precentral gyrus, including primary motor (BA4), somato-sensory (BA1-BA3), and somato-sensory association cortices (BA5), extending into the superior parietal lobule (BA7). Parietal SUVR localized dorsal to the intraparietal sulcus and lateral to precuneus.

Minimal SUVR localized to the left insula (BA13).

Pattern 7: maximal SUVR was bilateral, symmetric, and covered the rostral frontal lobes, including anterior prefrontal (BA10), orbitofrontal (BA11), orbital inferior frontal (BA47/12), and dorsolateral prefrontal cortical (BA46) areas. SUVR extended onto the medial surfaces of the anterior prefrontal and orbitofrontal cortices. Minimal SUVR localized to striatum and ventral thalamus.

Pattern 8: maximal SUVR was bilateral, symmetric, and covered dorsal frontal and parietal areas, including dorsomedial and dorsolateral portions of dorsolateral prefrontal cortex (BA9), frontal eye field (BA8), supplementary motor area (BA6), and primary motor cortex (BA4).

SUVR tapered into superior portions of the extrastriate cortex for visual association (BA19).

Pattern 9: maximual SUVR was bilateral, symmetric, and covered contiguous cortical surfaces of the dorsal cerebrum. The inferior margin of SUVR enclosed superior aspects of middle frontal gyri and superior lateral frontal gyri (lateral aspects of BA9, BA8, BA6), and superior parietal lobules. SUVR was absent throughout medial cortical surfaces. Minimal SUVR localized to striatum and dorsolateral thalamus.

Pattern 10: maximal SUVR ranged across the right lateral hemisphere, ranging from the triangular part of the inferior frontal gyrus (BA45), to the pars opercularis (BA44), the subcentral area (BA43), the supramarginal gyrus (BA40), and the angular gyrus (BA39). Tapering SUVR minimally localizes to posterior aspects of primary auditory cortex (BA41).

Pattern 11: maximal SUVR was bilateral, symmetric, and localized to regions of the straight and transverse venous sinuses.

Pattern 12: maximal SUVR was bilateral, symmetric, and encompassed large, contiguous surfaces of the cerebrum and cerebellum along their anterior, lateral and posterior aspects. The superior margin of SUVR enclosed the frontal poles and inferior aspects of the middle frontal gyri, extending caudally to also enclose the inferior parietal lobules as well as the parieto-occipital junction. Notably, the inferior margin of SUVR excluded subgenual cortex (BA25), entorhinal and perirhinal cortices (BA34, BA35, BA28). SUVR was minimal along medial cortical surfaces, with some localization to dorsal aspects of the cingulate cortex (BA31).

Minimal SUVR also localized to patches within striatum and posteromedial thalamus.

Pattern 13: maximal SUVR was bilateral, symmetric, and ranged over medial aspects of orbitofrontal cortex (BA11) and medial prefrontal cortex (BA10) with extensions into adjacent deep white matter. Maximal SUVR also localized to large regions incorporating striatum, thalamus, and midbrain.

Pattern 14: maximal SUVR was bilateral, symmetric, and ranged over cerebellar cortex inferior to locations of the transverse venous sinuses. Minimal SUVR localized to patches along the lateral aspects of the inferior temporal gyri and along orbitofrontal areas.

Pattern 15: maximal SUVR was midline, encompassing the walls of the quadrigeminal cistern and interpeduncular cistern. Moderate SUVR encompassed the anterior midbrain and the walls of the third ventricle.

Pattern 16: maximal SUVR favored the right hemisphere with moderate SUVR symmetrically localized in the left hemisphere. Maximal SUVR ranged along the inferior parietal lobule, over the angular gyrus (BA39) and into posterior aspects of the superior temporal gyri (BA22, BA41, BA42). Minimal SUVR localized to posterior surfaces of the precuneus, bilaterally (BA7).

Pattern 17: maximal SUVR was largely midline, encompassing primary and secondary visual cortices (BA17, BA18). Minimal SUVR localized to patches of the left superior temporal gyrus.

Pattern 18: maximal SUVR was bilateral and symmetric, ranging over the grey-white junction and deeper centrum semiovale underneath primary, secondary, and association visual cortices (BA17-BA19). Minimal SUVR extended into deep white matter along the medial surfaces of the lateral ventricles, and also extended into the deep white matter beneath the left posterior-superior temporal gyri and beneath the left marginal gyrus.

Pattern 19: maximal SUVR was midline, ranging over the posterior cingulate cortext (BA31) and precuneus (medial BA7). Minimal SUVR localized to the angular gyri, striatum, and thalamus bilaterally (BA 39).

Pattern 20: maximal SUVR favored the right superior temporal gyrus (BA22, BA41, BA42) and right supramarginal gyrus (BA40). Moderate SUVR localized to the left superior temporal gyrus (BA22) and left supramarginal gyrus (BA40).

Pattern 21 (1): maximal SUVR extended throughout the centrum semiovale, internal capsule, and globus pallidus, but did not involve hippocampal structures. Minimal SUVR localized to anteromedial thalamus and cerebellar white matter.

Pattern 22: maximal SUVR was bilateral, symmetric, and ranged over the superior parietal lobule (BA7). Minimal SUVR localized to the posterior thalamus and scattered patches of deep white matter.

Pattern 23: maximal SUVR was bilateral, symmetric, and ranged confluently over the orbital frontal cortices (BA11), extending into medial prefrontal areas (BA10). Minimal SUVR extended into insular cortex bilaterally (BA16).

Pattern 24: maximal SUVR was bilateral, symmetric, and ranged over all cingulate cortices (BA31 – BA33, BA23, BA24), extending into corpus callosum and adjacent white matter. Moderate SUVR localized symmetrically into striatum, thalamus, and white matter tracts deep within temporal and parietal lobes.

### Patterns of Covariance for Metabolism Are Distinct from Other Known Networks

Pattern 1, expressing the greatest glucose metabolism, was left-hemisphere dominant, and revealed topography encompassing the cortical areas commonly ascribed to language functions by historical lesion studies, by neurosurgical functional studies, and by task as well as resting fMRI. Pattern 1, however, encompassed more than Broca’s or Wernicke’s areas, and more than ventral attention network topography from resting fMRI, notably including also bilateral precuneus. Pattern 2 symmetrically revealed the anatomy of the insular cortex and opercular cortex, and indicated associations with medial prefrontal cortex, an association previously not observed in other neuroimaging studies. Pattern 3 symmetrically revealed an expansive topography of medial cortical regions of import for cognition and behavior, involving lower extremity motor function with numerous association areas, including dorsolateral prefrontal cortex and precuneus. Pattern 4, encompassing entorhinal cortices, amygdala, and hippocampi, has drawn much scrutiny in dementia research for its known roles in memory and mood regulation, but pattern 4 is notably segregated from the larger topography ascribed to the default mode of large-scale functionality. Pattern 4, in union with the orbital frontal topography of pattern 23, which demonstrated much lower aggregate glucose metabolism, reproduced one of the patterns of the 12-pattern model (Figure 3). Similar correspondences between models with varying spatial coarse-graining indicate the reproducibility of NMF PoC and support the hierarchical organization of brain function^21^ that has been observed elsewhere^22–24^. Patterns 5, 11, 14, and 15 segregate the cerebellum, known to have detailed connectivity to the canonical resting-state networks, but these NMF PoCs have not indicated similar topographies in FDG PET, likely reflecting the limitations of resolution of ADNI PET processing, but notable for assignment of three cerebellar patterns. At present, there are no criteria for over-fitting that can exclude the separations of patterns 5, 11, 14, and 15. While some patterns likely captured variability in extra-axial cerebrospinal fluid or atrophy, patterns 9 and 12, gross misregistration was excluded by detailed examination and visualization of spatially normalized imaging. Pattern 21 identified the centrum semiovale with striatum, not previously identified in neuroimaging studies, to our best knowledge. Remaining patterns have analogs in large-scale functional studies, but these remaining patterns from NMF are more compact and contiguous than those familiar from fMRI.

**Figure 3.**
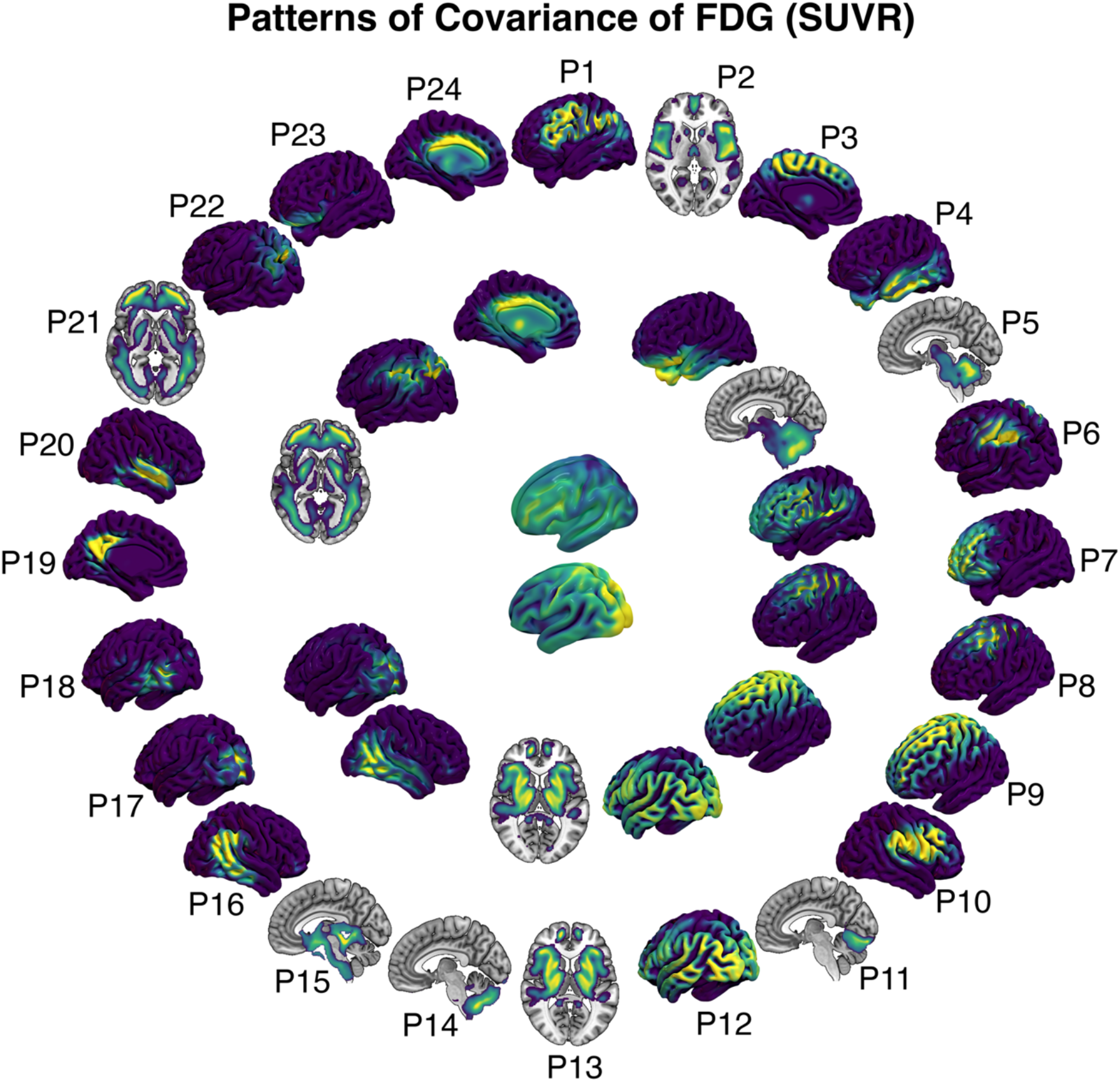
Patterns of covariance determined by model selection had peak selection objectives for 24 patterns, but 12 and 2 patterns provided local peaks of selection objective. Twelve patterns provided coarser grained patterns that nevertheless replicated patterns from the 24-pattern model. Two patterns provided regional segregation into analogs of the intrinsic (default mode and frontoparietal) and extrinsic (somatomotor sensory, auditory, visual) large-scale functional networks. The 2-pattern model is represented on a smoothed cortical surface for clarity of topographies.

### Effects of Age, Sex, APOE ε4

Age and sex were previously found significant for structural PoC by NMF in adolescent brain development^12^. GAMs for PoCs used *FDG ∼ s(age, k=20, interaction=sex) + sex + APOE*ε*4 + cohort,* following Wilkinson’s notation and denoting thin-plate regression splines with *s*, number of spline knots with *k*, and representing diagnostic cohorts with *cohort.* Categorical cohorts described contrasts compared to cognitively normal participants with CDR=0 and without amyloidosis. PoC SUVR decreased with age for CDR=0 without amyloidosis, but increased with age for patterns 16, 21, and for males with pattern 22, as shown in Figure 4. PoC SUVR decreased with age for CDR=0 with amyloidosis, but increased with age for females with pattern 16, for pattern 21, for males with pattern 22, and for females with pattern 23. PoC SUVR decreased with age for CDR=0.5 with amyloidosis, but increased with age for females with pattern 21 and females with pattern 22. Some male PoC had trajectories with age that were convex (PoC increased then diminished with age). PoC SUVR decreased with age for CDR>0.5 with amyloidosis, but increased with age for patterns 1 – 3, 6, 7, 11, 13, 16, 21, 22, and for females pattern 17, as shown in Figure 5. For the cognitively impaired cohort, Increasing glucose metabolism with age may indicate counter-regulatory activities in the presence of disease, but the role of glucose metabolism for the progression of dementia is also possible ^25,26^

**Figure 4.**
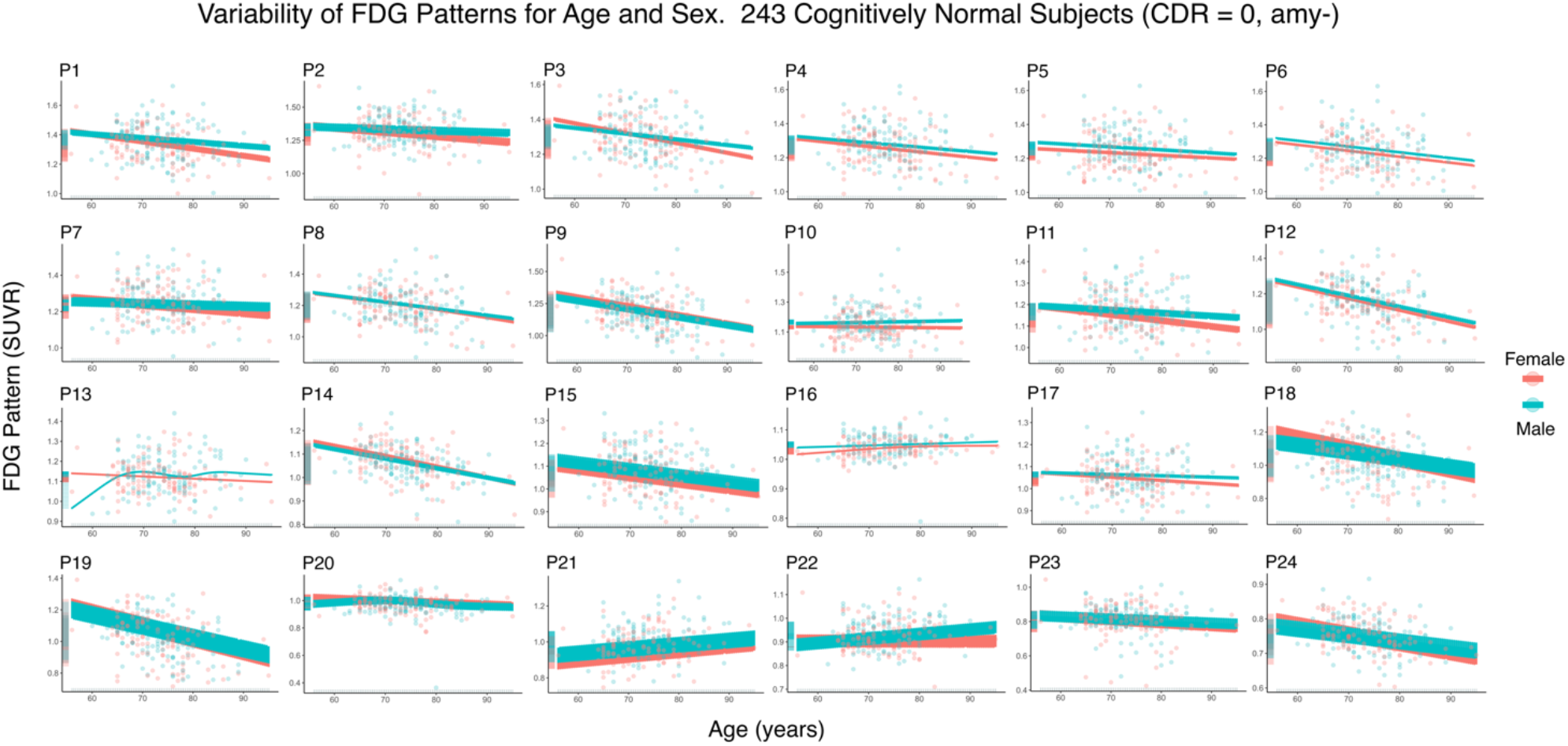
Generalized additive model of multivariate regression of patterns of covariance. FDG (SUVR) ∼ s(age, interaction=sex) + sex + apoe4 + cohort. The cohort with CDR=0 and no amyloidosis is shown. Confidence intervals are modulated by APOE ε4. FDG (SUVR) increases with age for P16, P21, and P22.

**Figure 5.**
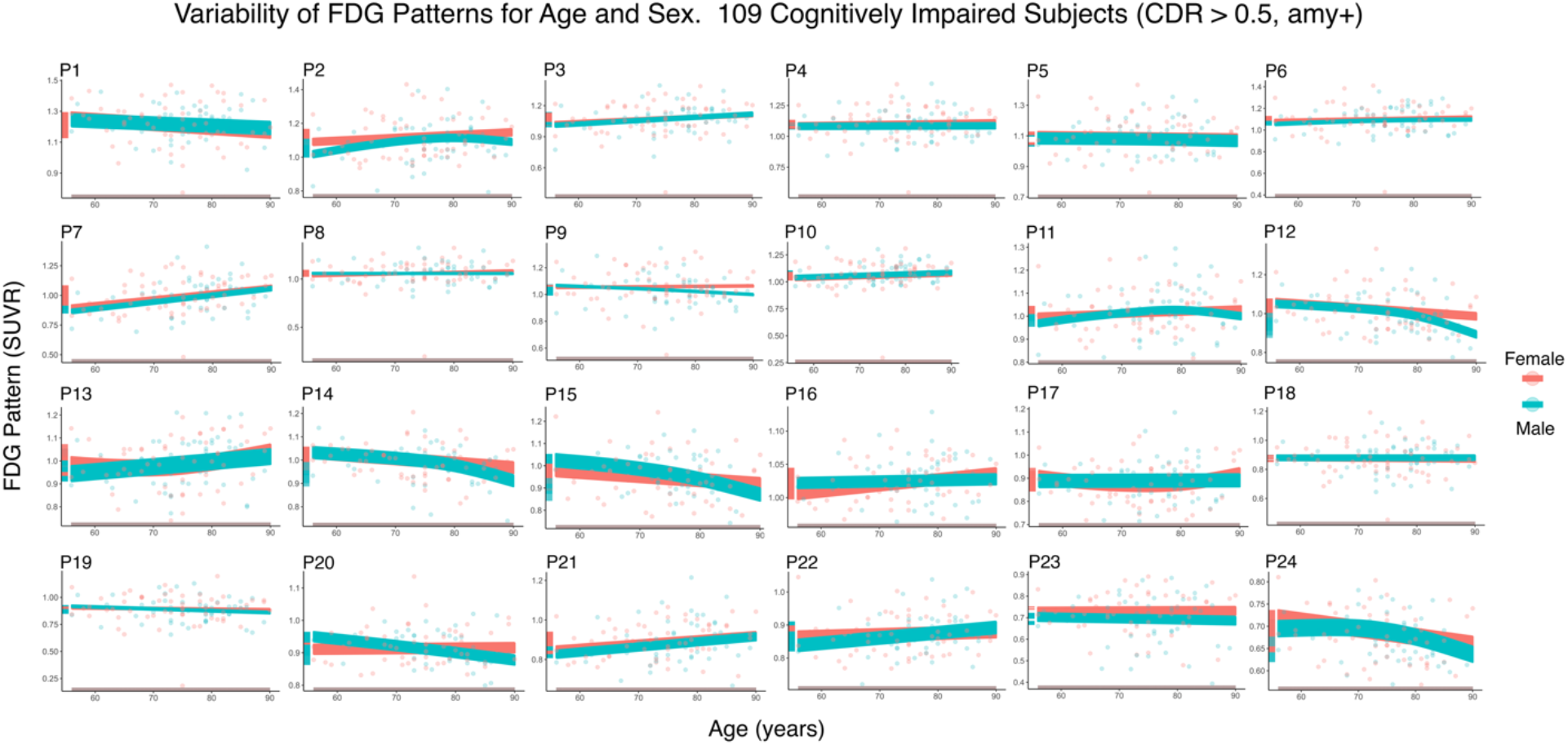
Generalized additive model of multivariate regression of patterns of covariance. FDG (SUVR) ∼ s(age, interaction=sex) + sex + apoe4 + cohort. The cohort with CDR=0 and no amyloidosis is shown. Confidence intervals are modulated by APOE ε4. FDG (SUVR) increases with age for P1 – P3, P6, P7, P11, P13, P16, P21, P22, and for females pattern P17. Symptomatic cohorts revealed increasing nonlinearities of FDG (SUVR) with age.

### Patterns of Covariance Indicate Neurodegeneration

Figure 6 shows plots of GAM coefficients relating diagnostic cohorts to the asymptomatic cohort without amyloidosis (CDR=0, amy-). GAM coefficients contrasting CDR=0, amy+ against CDR=0, amy-exhibited positivity, β _CDR=0,amy+_ > 0, for patterns 1, 8, 9, and 18 – 20, indicating increases of glucose metabolism for asymptomatic amyloidosis. However, the contrasts were not significant for our computed GAMs at the significance level of 0.05. Nevertheless, this positive constrast of glucose metabolism is consistent with observations of persistently youthful measures of aerobic glycolysis estimated from [^15^O]carbon-monoxide, [^15^O]oxygen, [^15^O]water, and FDG^26^. GAM coefficients contrasting CDR>0.5, amy+ against CDR=0, amy-, the contrasts for the severest cognitive impairments, were most negative for patterns 3, 2, and 7, illustrated in Figure 7.

**Figure 6.**
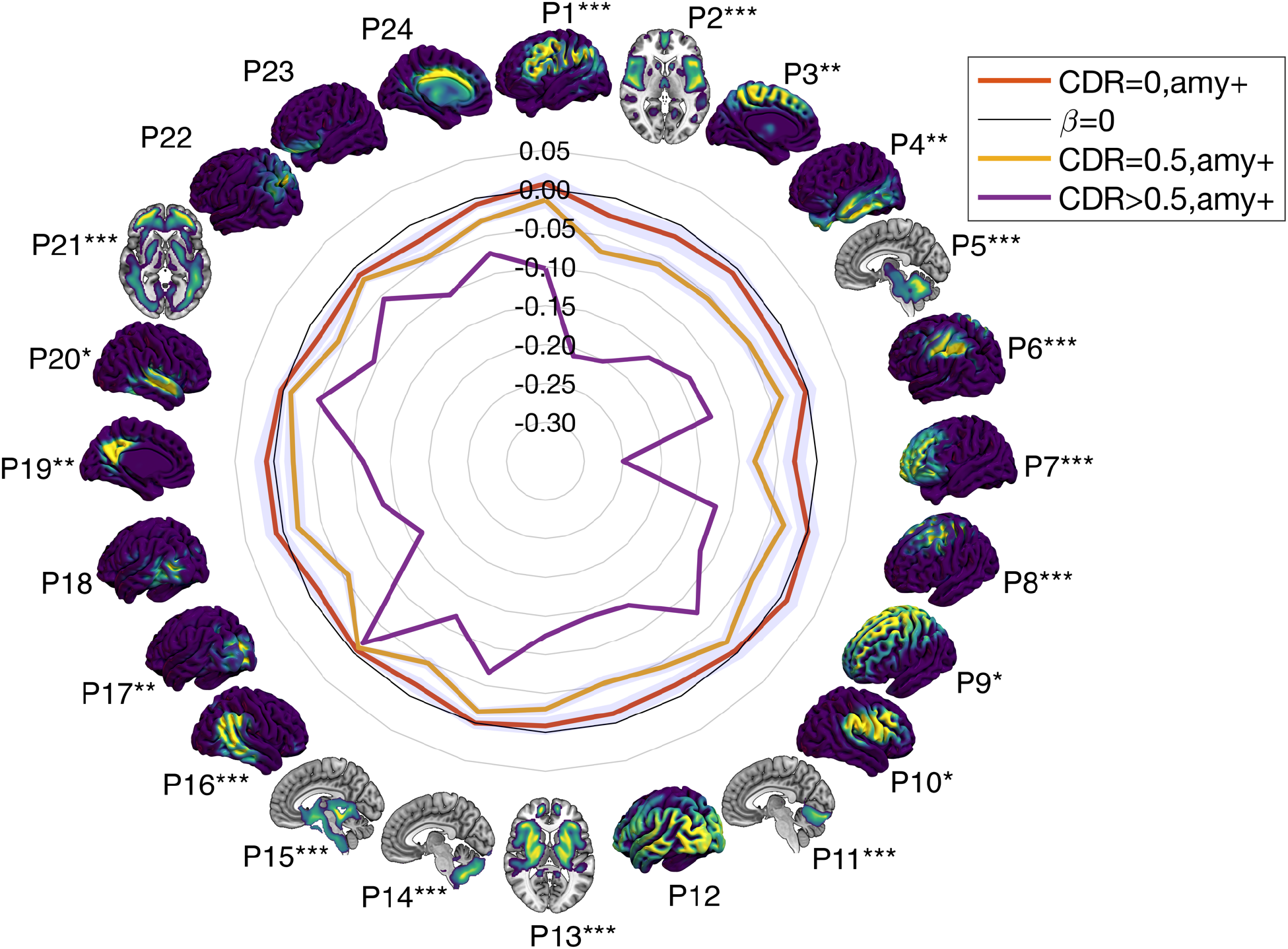
Generalized additive model of multivariate regression of patterns of covariance. FDG (SUVR) ∼ s(age, interaction=sex) + sex + apoe4 + cohort. Regression coefficients for diagnostic cohorts (CDR=0, amy+; CDR=0.5, amy+; CDR>0.5, amy+). The zero value for coefficients is indicated in gray. Coefficients describe variations of cohorts away from asymptomatic individuals without amyloidosis (CDR=0, amy-). Asterisks indicate p-values for coefficients of CDR=0.5, amy+, following Benjamini-Hochberg adjustments for false discovery rate: p < 0.05 ∼ *, p < 0.01 ∼ **, p < 0.001 ∼ ***. For asymptomatic individuals with amyloidosis (CDR=0, amy+), only pattern 18 was significant with p < 0.05. For moderately and severely symptomatic individual with amyloidosis (CDR>0.5, amy+), all patterns were significant to p < 0.001.

**Figure 7.**
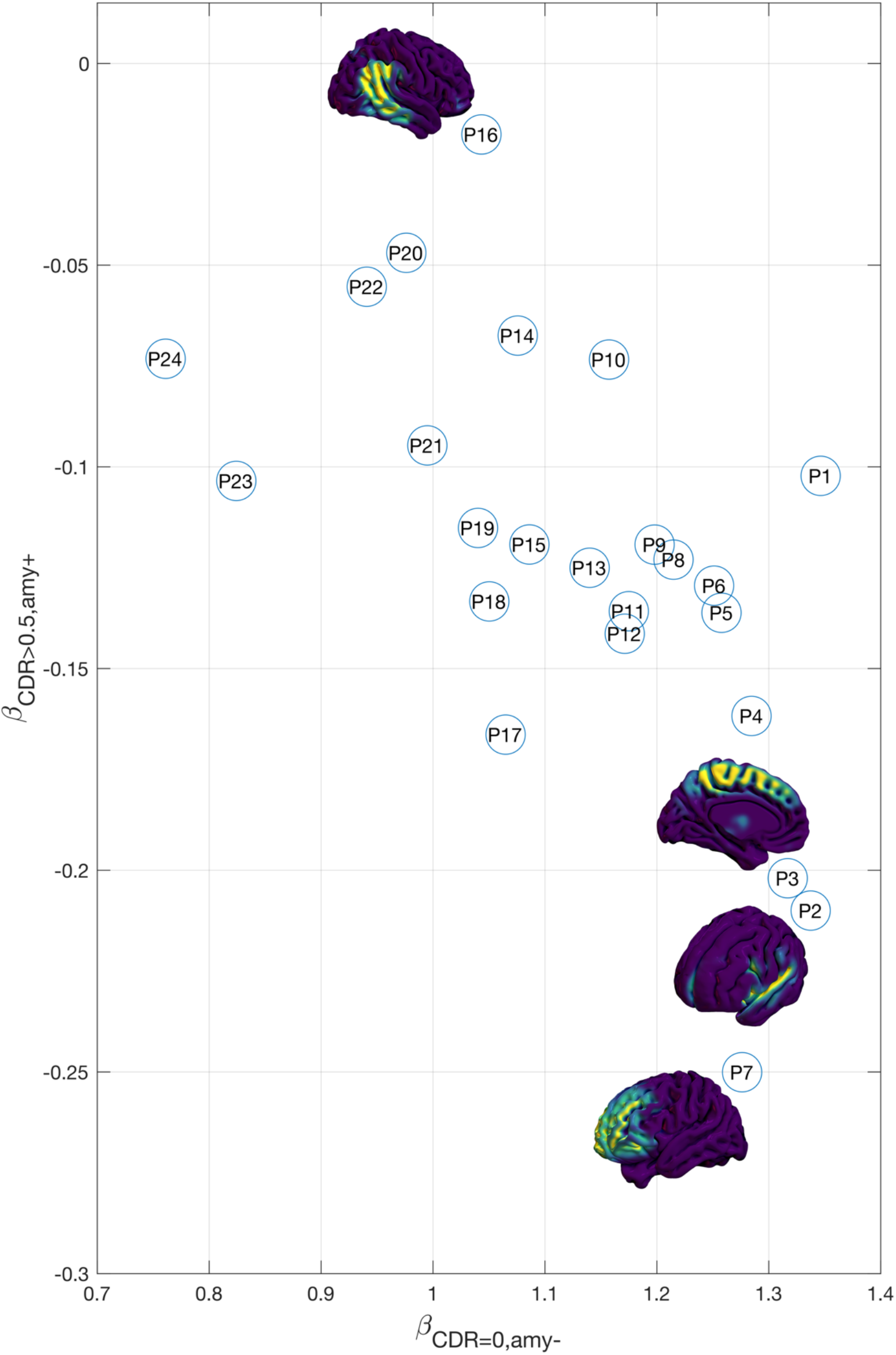
Patterns of covariance illustrated with GAM multivariate regression coefficients for the cognitively normal cohort (CDR=0, amy-) and severely cognitively impaired cohort (CDR>0.5, amy+). Patterns 7, 2, and 3 corresponded to the greatest loss of glucose metabolism, and inferred neurodegeneration with progression of disease. Pattern 16 retained glucose metabolism even for the severely cognitively impaired. All coefficients had p-values < 0.0001.

Remarkably, for all constrasting GAM coefficients for cohorts, pattern 16 was largely preserved, also illustrated in Figure 7.

### Predictive Patterns of Neurodegeneration Correspond to Cognitive Domains

Figure 7 clarifies that neurodegeneration corresponds to losses of glucose metabolism along the cortical midline, within the lateral sulcus, insula, medial prefrontal cortex, and the frontal pole. These cortical regions, identified as PoC 3, 2, and 7, have larger coefficients in multivariate generalize linear modeling (GLM) associating them to dependent variable for Alzheimer’s dementia, especially CDR, CDR-SOB, and metrics of tau. Multivariate GLM results are summarized in Figure 8. Most PoCs had sex dependence, as anticipated from existing literature on cognitive impairment and glucose metabolism^27^. In multivariate GLM, APOE ε2 was also broadly dependent upon PoC.

**Figure 8.**
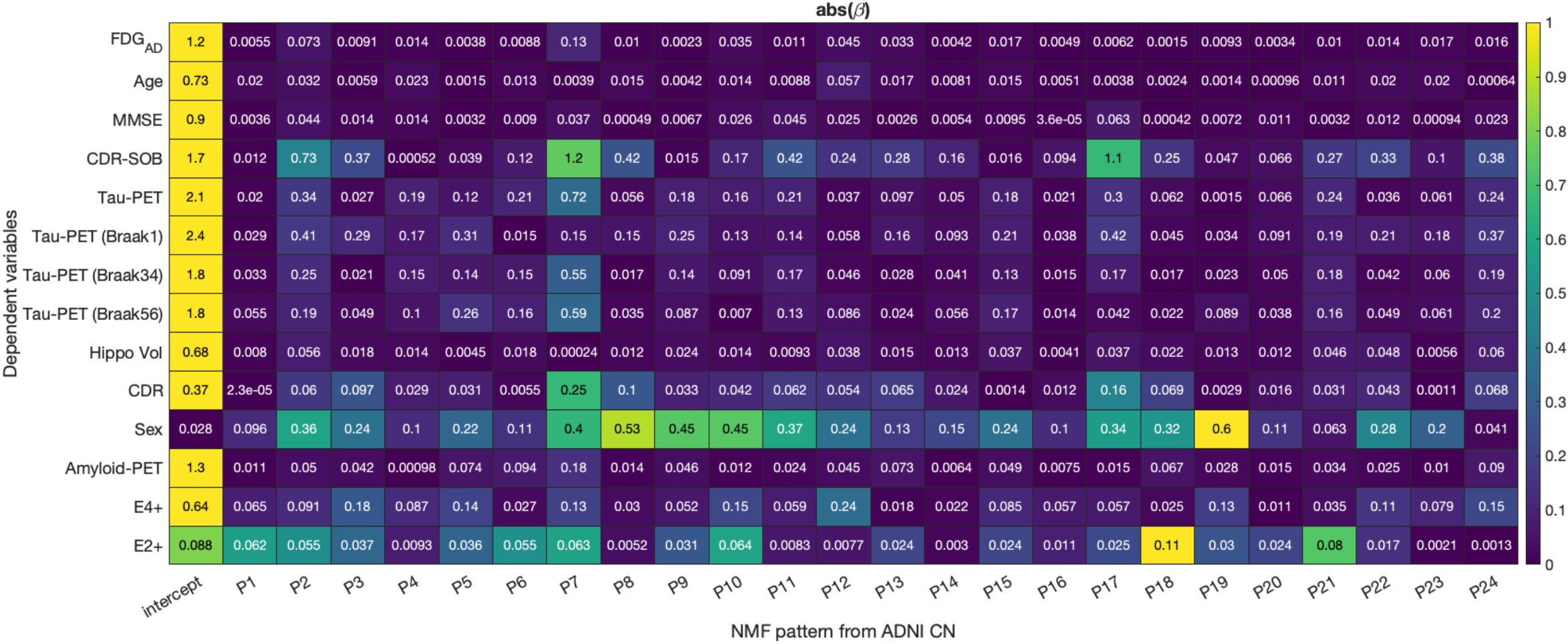
Multivariate generalized linear model for patterns of covariance and dependent variables relevant for Alzheimer’s dementia. FDG_AD_ are SUVR in Alzheimer’s disease signature regions. MMSE is the Mini-Mental State Examination. CDR-SOB is Clinical Dementia Rating Scale Sum of Boxes. Tau-PET are SUVR from PET with tau-binding tracers. Tau-PET (Braak1, Braak34, Braak56) are staging scores from post-mortem neurofibrillary tangle estimates. Hippo Vol is hippocampal volume. CDR is Clinical Dementia Rating Scale. Amyloid-PET are SUVR from PET with amyloid-binding tracers. E4+ indicates carriage of APOE ε4 alleles. E4+ indicates carriage of APOE ε2 alleles.

Remarkably, neurodegeneration and glucose metabolism were preserved in the right angular gyrus, supramarginal gyrus, and posterior aspects of the superior and middle temporal gyri, especially posterior to Heschl’s gyrus, PoC 16, shown in Figure 7.

Utilizing spin-testing of inflated cortical surface^28^ and corrections of volumetric auto-correlations^29^, we mapped PoCs of glucose metabolism to topical terms from the Neurosynth platform^9,30,31^. This provided commonly semantic decodings comparable to encodings made for fMRI^20,32^ and previous studies of FDG PET^2^. These mappings for 104 topic terms previously curated for statistical independence^28^ are illustrated in Figure 9. A graph of PoC with significant correlations with topic terms is shown in Figure 10.

**Figure 9.**
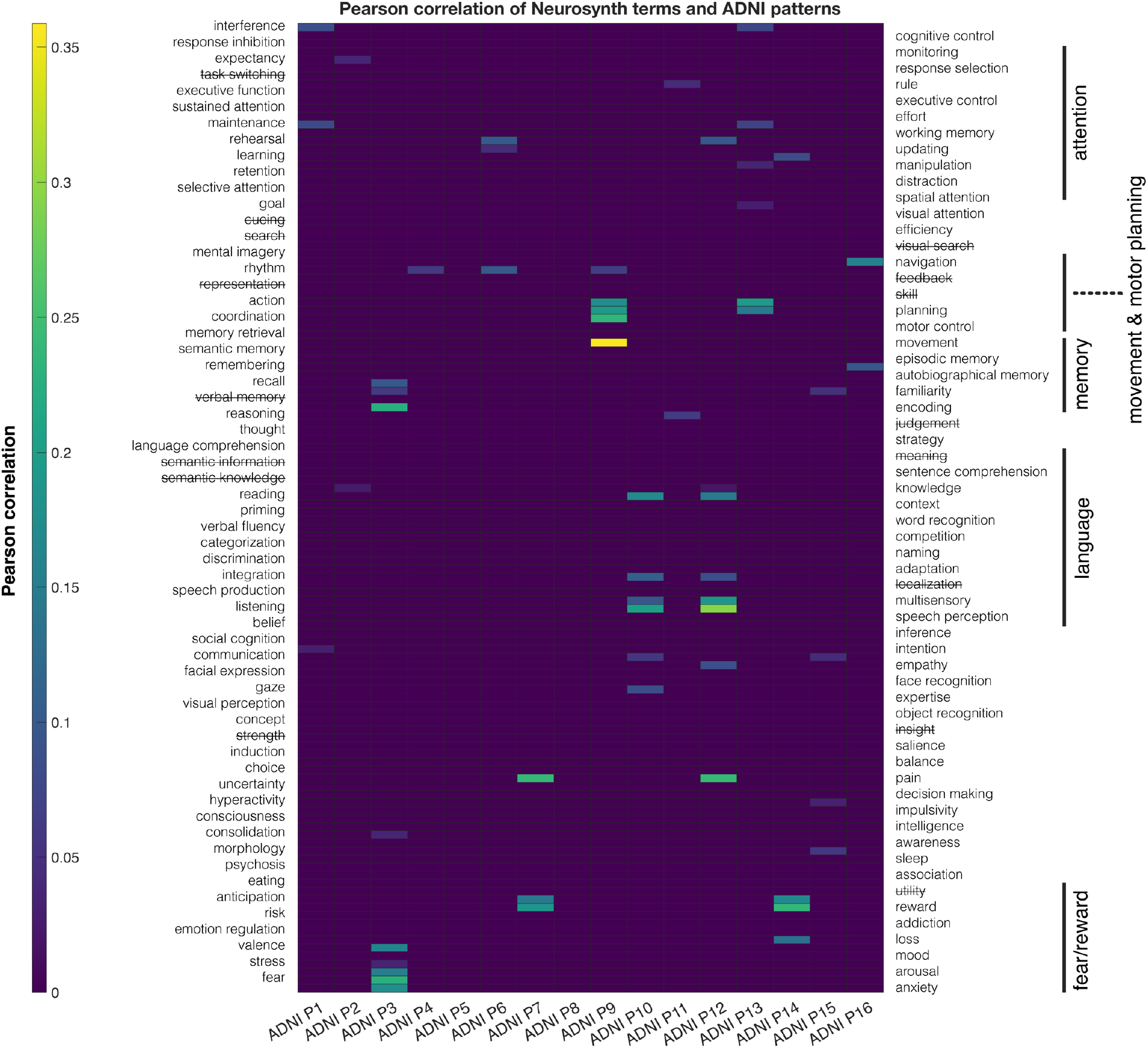
Pearson correlations of patterns of covariance with Neurosynth topical terms selected by spin-testing (Alexander-Bloch et al. 2018). The NMF model with 16 components is illustrated. Significant correlations were identified by correction of volumetric spatial autocorrelations using the framework of Brainsmash (Burt et al., 2020).

**Figure 10.**
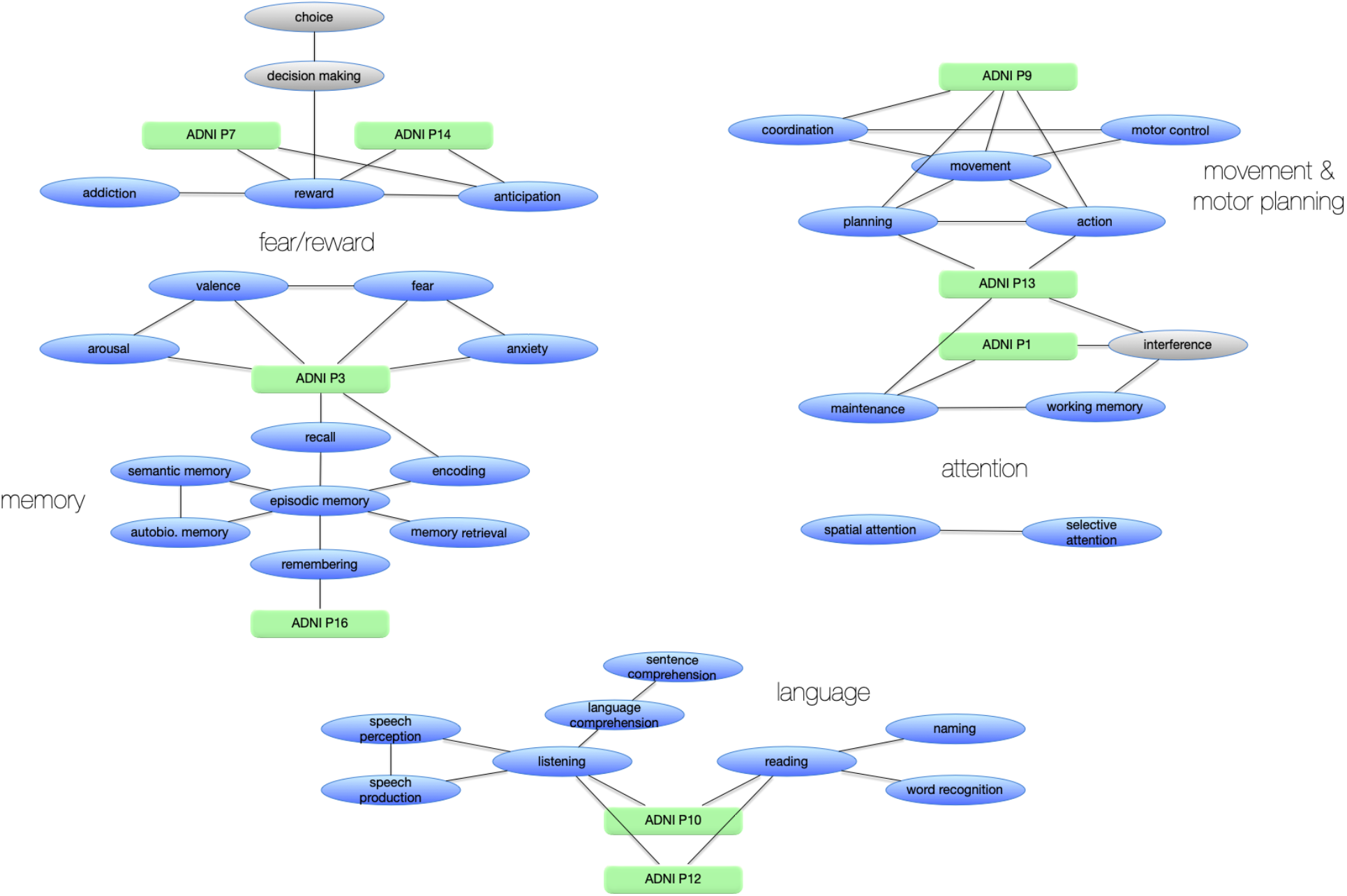
Patterns of covariance are embedded in graphs of significantly connected topical terms from Neurosynth.

## Discussion

This study used advanced multivariate methods to make retrospective inferences on Alzheimer’s disease in ADNI, a large, multicenter, deeply curated, publicly accessible repository of clinical and neuroimaging data. The primary study result is a collection of PoC that describe factors ascribable to regional glucose metabolism as estimated from FDG SUVR^14^ using NMF^12,33^. The construction of PoC using NMF on a well-defined cognitively normal cohort was data-driven, without a priori hypotheses concerning ADNI data^34^, following methods for NMF previously used successfully for neuroimaging inferences^12,35^. This study examined the multivariate covariances of the NMF PoC with well-known variables for the progression of neurodegeneration to Alzheimer’s disease, including cortical topography^32^, age, sex^27^, and APOE ε4 alleles^36^. PoC constructed from the cognitively normal cohort provided spatially distributed bases for inference of cohorts with progressive CDR in the presence of cerebral amyloidosis. GAMs^37,38^ demonstrated that nonlinear dependence on age, interactions with sex, and influence of APOE ε4became more prominent with progressive neurodegeneration. Neurodegeneration was especially sizeable for PoC encompassing insular cortex, frontal cortex and midline frontoparietal cortical surfaces. Notably, neurodegeneration spared lateral parietal and posterior superior temporal areas, even in severely symptomatic ADNI participants with amyloidosis. In asymptomatic ADNI participants, PoC in selective areas demonstrated increasing glucose metabolism in the presence of amyloidosis compared to those without amyloidosis, reproducing recent observations by Goyal et al. for aerobic glycolysis for which oxygen metabolism was minimally contributory^26^.

### Topography of Glucose Metabolism and Neurodegeneration

PoC were reproducible at multiple scales (number of model patterns). The degeneration of the default mode network in Alzheimer’s disease has been understood for decades^3^, but PoC from this work demonstrated that more granular topographies characterized neurodegeneration. This work on FDG PET reiterated^12^ four keys aspects for PoCs constructed from NMF. First, higher PoC resolutions respected boundaries of lower resolutions, consistent with hierarchical organizations of patterns. Without constraints favoring spatial contiguity or sparsity, PoC from NMF decompositions were contiguous and sparse, supporting small world properties^23,24^. There were no constraints favoring spatial positions or symmetries, yet PoC from NMF retained most neurostructural symmetries. PoC also revealed expected asymmetries of ventral attention or language functionality. Modest asymmetries emerged in pattern 16, suggesting avenues for future investigations of brain regions that are resilient to neurodegeneration. PoC revealed associations that crossed gyral anatomy, consistent with microarchitectural and functional priors. However, PoC associations were distinct from known atlases of structure and function, indicating uniqueness of regional glucose metabolism. PoC revealed overlapping areas, consistent with concurrently distributed underlying processes driving glucose metabolism.

Neurosynth provided semantic mappings in accord with previous approaches with eigenbrains^2^, and yielded novel features that may improve models of neurodegeneration. Fear and reward semantics were coded by PoCs for insula, cingulate cortices, and medial prefrontal cortex. Memory was coded by PoCs for inferior temporal (entorhinal), posterior cingulate, and precuneal areas as expected, with addition of secondary and associative visual cortices. The latter have historically associated with dementia variants affecting vision in younger cohorts. Language was coded by PoCs for lateral parietal, posterior temporal, opercular and insular areas, largely as expected from familiar functional topographies. Movement and motor planning was coded by PoCs for somato-motor-sensory cortex, premotor areas, and secondary motor areas, but also deeper white matter.

### Limitations

This work is retrospective, using a mature dataset which has been studied from numerous previous scientific perspectives since the first public availability of ADNI data. While benefitting from breadth and depth of curation, public datasets can accumulate implicit biases. However, we are unaware of any known biases that could affect this work. This work makes inferences on cross-sectional data corresponding to the time of enrollment of participants in ADNI, which also may incur biases of selection and timing. For the date ranges chosen in this work, 1165 subjects accumulated 1890 imaging sessions with FDG PET, from which modest longitudinal inferences could be made. Nevertheless, the multivariate strategy of the analyses brings forth contrasts among differing brain regions, reducing the influence of data collections made at baseline epochs. The generalizability of NMF PoC derived from FDG to external, unseen data will be tested in follow-up studies of replication.

### Study Implications

The resulting PoC from this study may provide important alternative regional criteria, distinct from atlas regions, and made specific for glucose metabolism which has known pathophysiologic mechanisms corresponding to neurodegeneration. In the pursuit of high-dimensional datasets, specific regional PoC for neurodegeneration will provide rational means of dimensionality reduction for purpose of interpretability and predictions. PoC results from this study can be reused in combinations with other neuroimaging data, with full use of computational automations, to achieve models with greater predictive capabilities.

### Conclusions

NMF is a data-driven, principled, supervised statistical learning method that provides interpretable patterns from neuroimaging. It can indicate regions specific for parts-based features that exhibit loss of glucose metabolism on FDG PET, thereby indicating neurodegeneration. It provides lower-dimensional models that can help inform the understanding and treatment of Alzheimer’s disease.

## Methods

*****

### Participants and Neuroimaging

#### ADNI data selection

Data for all participants were drawn directly from ADNI (https://adni.loni.usc.edu) and the ADNI Data Package for R (The ADNI Team. ADNIMERGE: Alzheimer’s Disease Neuroimaging Initiative, R package version 0.0.1 (2023); https://adni.bitbucket.io/index.html). Following filing of data use agreements, all data were downloaded via ADNI’s web services for data access. FDG PET were primary data objects and data curation began with collection of all available FDG PET from 9/22/2005 – 1/4/2022 which were co-registered, averaged, standardized for image and voxel size, and transformed to uniform resolution. For each subject with FDG PET, subsequent data gathering included: all available T1-weighted (T1w) imaging, and all available dataframes for ADNIMERGE in comma-separated-value formats. T1w imaging provided native anatomy for anatomical inferences and for nonlinear spatial normalizations. For each FDG PET, we selected the most contemporaneous T1w imaging, not exceeding 365 days separation from FDG PET. We curated dataframe values so as to ensure that all values used for inferences were contemporaneous to within 365 days. We gathered scalar FDG SUVR from meta-ROIs, age, MMSE, CDR-SOB, scalar tau-specific SUVR, Braak staging measures, hippocampal volume, CDR, sex, amyloid-specific SUVR, APOE ε4, and APOE ε2. Ultimately, we excluded from inferences any FDG PET that lacked adequate contemporaneous ancillary data.

#### Image-processing pipelines

All imaging was nonlinearly warped to the MNI152 atlas. First, we ensured all imaging to be formatted to NIfTI using dcm2niix^39^. Then, we corrected bias fields in T1w imaging using ANTs N4BiasFieldCorrection^40,41^. Next, we constructed binary masks for the whole brain using DeepMRSeg^42^. Next, we constructed nonlinear warps using, from ANTs, antsRegistrationSyNQuick and antsApplyTransforms. Next, using whole brain masks, we used 4dfp t4_resolve (https://4dfp.readthedocs.io) to obtain rigid-body co-registration of FDG to T1w images. Next, we adjusted normalizations of FDG SUVR to values in FreeSurfer-determined pons and cerebellar vermis^15^. Composition of all registrations, warpings, and their inverses, produced transformations for FDG onto the MNI152 atlas. Finally, we applied the atlas-registered binary mask to exclude all FDG voxels not in the brain, voxels which otherwise were needed for high-resolution warping.

The final quality assurance procedure used human visualization of all FDG imaging providing for construction of PoC by NMF. Greatest variability appeared in the posterior cerebellum, the cerebral vertex in the vicinity of the sagittal sinus, and cortical thickness after nonlinear registration. Therefore, NMF patterns involving these regions likely represent aspects of nonlinear misregistration.

### Multivariate Analysis of Hierarchical Covariance Structures

To infer hierarchical covariance structures in ADNI participants and their FDG PET, we used NMF. FDG PET from all participants (*n* in number) require reshaping such that all voxels from a single PET session (*d* in number) comprise a column vector, and horizontal concatenation of vectors from all participants form a data matrix, 𝑋 = [𝑥_1_, …, 𝑥_𝑛_], 𝑥_i_ ∈ 𝑅^𝑑^, of size *d* x *n*. All PET voxels are non-negative after reconstruction of emission activities. Consequently, the sought approximate factorization is 𝑋 ≈ 𝑊𝐻 with 𝑊 = [𝑤_1_, …, 𝑤_𝑘_], 𝑤_i_ ∈ 𝑅^𝑑^ and 𝐻^𝑇^ = [ℎ_1_, …, ℎ_𝑘_], ℎ_i_ ∈ 𝑅^𝑛^, with all elements of *W* and *H* also being non-negative. The number of adjustable patterns 𝑘 ≪ 𝑑 and 𝑘 ≪ 𝑛, thereby providing dimensionality reduction. Matrix *W* provides a *d-*voxel representation for each of *k* patterns in each column. Matrix 𝐻^𝑇^provides an *n-*participant representation, describing the variability of the participants, for each of *k* patterns in each column. Our implementation of NMF (https://github.com/asotiras/brainparts) imposes constraints for orthonormality, 𝑊^𝑇^𝑊 = 𝐼, and for projection to participant representations, _𝐻 = 𝑊_𝑇_𝑋_^13,43^.

We ran multiple NMF trials for 2-40 patterns with which we performed model selection to find the optimal number of patterns consistent with our data. Split-half reproducibility made principled use of anticlustering to find minimally clustered splits^44^, then examined the stability of NMF results for each of the trials of patterns^45,46^. The optimal number of patterns satisfied reproducibility: results with the highest mean adjusted Rand index (ARI) across 49 split-half bootstraps; and reliability: results with the lowest deviation of ARI across bootstraps^47,48^. ARI measures set similarity adjusted for chance, allowing for balanced comparisons between sets of patterns for variable numbers of patterns. We also examined reconstruction errors as the Frobenius norm between data matrix, *X*, and the NMF decomposition, *WH*.

### Analysis of Patterns of Neurodegeneration

We accounted for spatial autocorrelations of volumetric FDG imaging as many features meaningful for the study were subcortical. For this purpose, we used BrainSMASH (https://github.com/murraylab/brainsmash)^29^.

### Estimation of Metabolic and Cognitive Associations

Finding ontological correspondence between the body of known neuropsychological landmarks and FDG imaging features required use of the Neurosynth platform (https://neurosynth.org)^9^.

## Data Availability

https://neurovault.org/collections/13302/

## Code Availability

https://github.com/jjleewustledu/mladni/tree/master

## Abbreviations

(ADNI): Alzheimer’s Disease Neuroimaging Initiative
(BA): Brodmann area
(CDR): clinical dementia rating
(FDG): fluorodeoxyglucose
(fMRI): functional magnetic resonance imaging
(GAM): generalized additive model
(GLM): generalized linear model
(MRI): magnetic resonance imaging
(NMF): nonnegative matrix factorization
(PET): positron emission tomography
(PCA): principal component analysis
(PoC): patterns of covariance
(SUVR): standardized uptake value ratio

## Acknowledgements

This work was supported by the National Institutes of Health (NIH) (R01-AG067103). Computations were performed using the facilities of the Washington University Research Computing and Informatics Facility, which were partially funded by NIH grants S10OD025200, 1S10RR022984-01A1 and 1S10OD018091-01. Additional support is provided The McDonnell Center for Systems Neuroscience.

Data collection and sharing for this project was funded by the Alzheimer’s Disease Neuroimaging Initiative (ADNI) (National Institutes of Health Grant U01 AG024904) and DOD ADNI (Department of Defense award number W81XWH-12-2-0012). ADNI is funded by the National Institute on Aging, the National Institute of Biomedical Imaging and Bioengineering, and through generous contributions from the following: AbbVie, Alzheimer’s Association; Alzheimer’s Drug Discovery Foundation; Araclon Biotech; BioClinica, Inc.; Biogen; Bristol-Myers Squibb Company; CereSpir, Inc.; Cogstate; Eisai Inc.; Elan Pharmaceuticals, Inc.; Eli Lilly and Company; EuroImmun; F. Hoffmann-La Roche Ltd and its affiliated company Genentech, Inc.; Fujirebio; GE Healthcare; IXICO Ltd.; Janssen Alzheimer Immunotherapy Research &Development, LLC.; Johnson & Johnson Pharmaceutical Research & Development, LLC; Lumosity; Lundbeck; Merck & Co., Inc.; Meso Scale Diagnostics, LLC.; NeuroRx Research; Neurotrack Technologies; Novartis Pharmaceuticals Corporation; Pfizer Inc.; Piramal Imaging; Servier; Takeda Pharmaceutical Company; and Transition Therapeutics. The Canadian Institutes of Health Research is providing funds to support ADNI clinical sites in Canada. Private sector contributions are facilitated by the Foundation for the National Institutes of Health (www.fnih.org). The grantee organization is the Northern California Institute for Research and Education, and the study is coordinated by the Alzheimer’s Therapeutic Research Institute at the University of Southern California. ADNI data are disseminated by the Laboratory for Neuro Imaging at the University of Southern California.

